# Four dimensional-flow magnetic resonance imaging analysis of carotid-cavernous fistula, dural arteriovenous fistula and spinal arteriovenous fistula: Detecting shunt point and diagnosing based on flow dynamics analysis

**DOI:** 10.1101/2023.12.05.23299574

**Authors:** Hiroto Kawano, Shigeki Yamada, Atsushi Tsuji, Takaaki Kubo, Akira Teshigahara, Shohei Ishida, Futa Ninomiya, Yuki Fujimoto, Tomoaki Kitamura, Satoshi Shitara, Yayoi Yoshimura, Kenji Takagi, Yoshiyuki Watanabe, Kazuhiko Nozaki, Kazumichi Yoshida

**Affiliations:** Department of Neurosurgery, Shiga University of Medical Science, Shiga, Japan; Department of Neurosurgery, Nagoya City University Graduate School of Medical Science, Aichi, Japan; Department of Neurosurgery, Omi Medical Center, Shiga, Japan; Department of Radiology, Shiga University of Medical Science, Shiga, Japan; Department of Neurosurgery, National Hospital Organization Higashi-Ohmi General Medical Center, Shiga, Japan

## Abstract

**Background and purpose:** Carotid-cavernous fistulas (CCF), dural arteriovenous fistula (DAVF) and spinal arteriovenous fistula (SAVF) are rare vascular malformations that cause arteriovenous shunting. Detecting the shunt point and flow direction is necessary for shunt disease diagnosis and treatment. Hence, we propose a new shunt disease diagnostic way with four dimensional-flow magnetic resonance imaging (4D-flow MRI).

**Material and methods:** The 4D-flow MRI and the DSA (digital subtraction angiography) were performed on 23 patients, including 2 with CCF, 20 with intracranial DAVF, and 1 with SAVF. Catheter embolization was performed in 19 patients. The 4D-flow MRI assessment was conducted for pre- and post-treatment (n = 9), pretreatment only (n = 12) and post-treatment only (n = 2). We use two functions of 4D-flow imaging application, one is a two-dimensional (2D)-colormap, which indicates velocity distribution with color scale for detecting shunt point, and another one is a 2D-flow vector for diagnosing flow direction.

**Results:** Shunt points are detected as red-colored lesions, with higher velocity compared with the surrounded area on 2D-colormap, with corresponding results to DSA findings. A 2D-flow vector indicates flow vectors of the main feeder, drainer, and influenced dural venous sinuses that could complement information obtained from other standard examinations. The post-therapeutic residual shunt flow analysis was consistent with DSA results except for one DAVF case.

**Conclusion:** 2D-colormap helps detect the shunt point of vascular malformations. A 2D-flow vector makes it possible to diagnose CCF, DAVF and SAVF based on flow dynamic analysis. A 4D-flow MRI would be an additional measurement for shunt disease diagnoses.

## Introduction

Advancements in MRI technique and three-dimensional (3D) medical imaging workstation allow us to use 4D-flow MRI for clinical practice of several diseases ^1–5^. A 4D-flow MRI is flow information in 3D space with a time axis, which consists of 3D morphological information and 3-directional phase imaging of flow information ^6,7^. Acquisition times of 4D-flow MRI with the appropriate spatial resolution is enough imaging duration for clinical practice application and obtain the needed information for blood vessel analysis minimally 1 mm in diameter.

CCF, DAVF and SAVF are arteriovenous shunt diseases that required an understanding of flow dynamics for pathological diagnosis and effective treatment ^8–12^. Various imaging measurements, including ultrasonography, contrast-enhanced computed tomography (CT), DSA, and MRI, have been used for evaluating CCF, DAVF and SAVF; however, past examinations were insufficient to diagnose from the perspective of precise flow dynamics ^13–18^. MRI development, as spatial resolution and time axis is one of the technical solutions for detecting flow information that had been unavailable by past standard examination methods. Our study is based on prior research of 4D-flow MRI for the evaluation of DAVF ^19^ ^20^.

This study aimed to indicate the feasibility of 4D-flow MRI to diagnose CCF, DAVF and SAVF based on a sufficient understanding of disease flow dynamics. The main theme of the study is shunt point and flow vector of shunt disease investigation by 4D-flow MRI.

## Methods

### Patient Characteristics

A total of 23 patients (13 males and 10 females), aged 16–80 years (mean age, 60.0 years) diagnosed with CCF, DAVF or SAVF underwent MRI examination from July 16, 2020, to October 24, 2023. All patients were symptomatic and diagnosed as CCF, DAVF or SAVF before performing a 4D-flow MRI. All cases were classified using the Barrow and Cognard classification ^21,22^ and classification for SAVF ^23^ ^24^, which revealed two direct CCF, with one Barrow type A and one type B; five cavernous sinuses (CS) DAVF, two cases were Cognard type I, one case was type IIa and two cases were type IIa+b; ten DAVF of the transverse sinus (TS) to sigmoid sinus (SS), one case was Cognard type I, two case was type IIa, three cases were type IIa+b, three cases were type III and one case was type IV; two DAVF of the anterior cranial fossa(ACF) of Cognard type IV; two DAVF of the sphenoid ridge(SR), one case was Cognard type III, and one case was Congnard type IV; one DAVF of the petrous apex(PA) of Cognard type IV; one SAVF case of conus perimedullary AVF. A total of 21 4D-flow MRIs and 21 DSAs for diagnosis and 19 DSA for treatment were performed. All patients underwent DSA for diagnosis and conventional MRI besides 4D-flow MRI. The assessment of 4D-flow MRI was conducted for pre-and post-treatment (n = 9), pretreatment only (n = 12), and post-treatment only (n = 2). Nineteen patients received endovascular treatment, including transvenous coil embolization with or without an intracranial stent and transarterial embolization using a platinum coil with or without *n*-BCA or Onyx (ev3, Irvine, California) injections (table.1). Two patients underwent endovascular therapy two times, therefore 3 times of 4D-flow MRI and DSA were executed and evaluated for them, respectively. One patient received endovascular therapy three times, two in other hospitals, and the patient underwent only a preoperative 4D-flow MRI assessment for retreatment. Preoperatively, 4D-flow MRI and DSA for diagnosis were consecutively performed on the same day or within a few days (mean, 2.9 days; range, 0–21 days). The time interval between the endovascular therapy and postoperative 4D-flow MRI was low (mean, 39.0 days; range, 1–194 days).

**Table 1.**
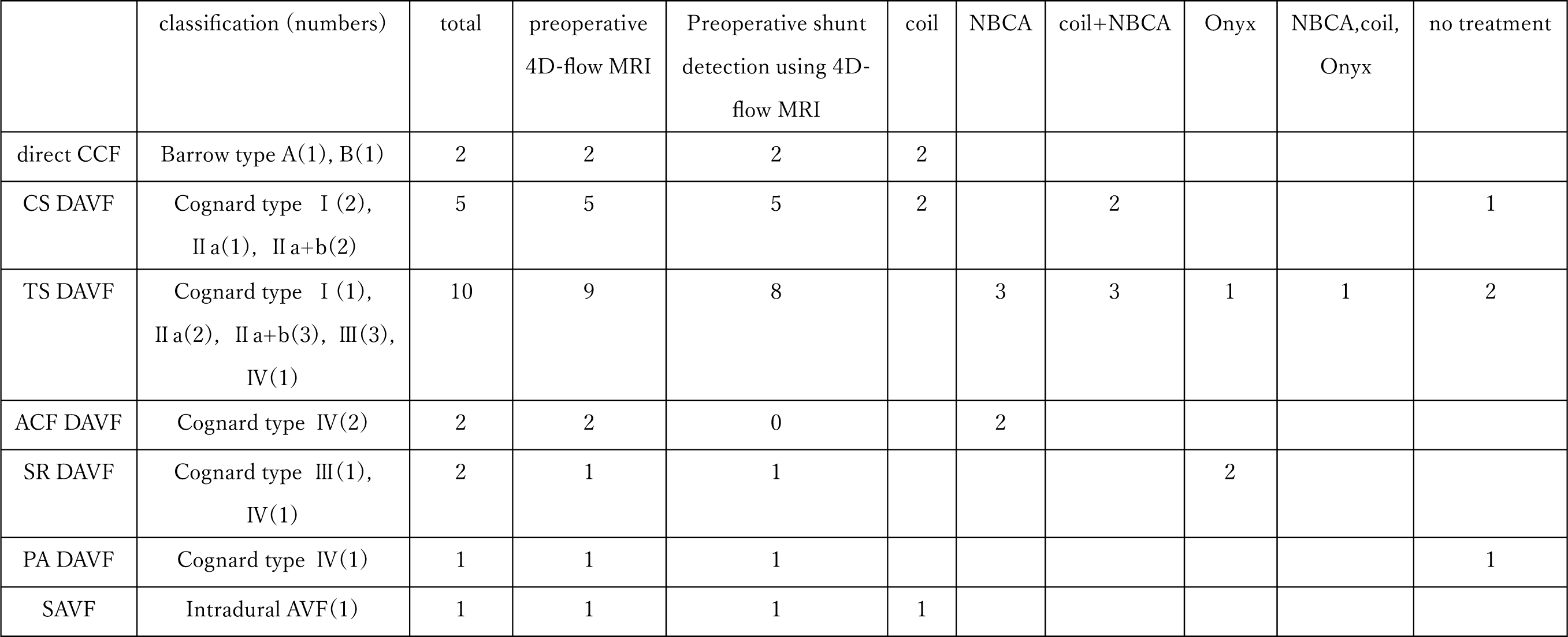
Summary of classification of shunt diseases, preoperative 4D-flow MRI and treatment.

|  | classification (numbers) | total | preoperative<br>4D-flow MRI | Preoperative shunt<br>detection using 4D-flow MRI | coil | NBCA | coil+NBCA | Onyx | NBCA,coil,<br>Onyx | no treatment |
| --- | --- | --- | --- | --- | --- | --- | --- | --- | --- | --- |
| direct CCF | Barrow type A(1), B(1) | 2 | 2 | 2 | 2 |  |  |  |  |  |
| CS DAVF | Cognard type I (2),<br>II a(1), II a+b(2) | 5 | 5 | 5 | 2 |  | 2 |  |  | 1 |
| TS DAVF | Cognard type I (1),<br>II a(2), II a+b(3), III(3),<br>IV(1) | 10 | 9 | 8 |  | 3 | 3 | 1 | 1 | 2 |
| ACF DAVF | Cognard type IV(2) | 2 | 2 | 0 |  | 2 |  |  |  |  |
| SR DAVF | Cognard type III(1),<br>IV(1) | 2 | 1 | 1 |  |  |  | 2 |  |  |
| PA DAVF | Cognard type IV(1) | 1 | 1 | 1 |  |  |  |  |  | 1 |
| SAVF | Intradural AVF(1) | 1 | 1 | 1 | 1 |  |  |  |  |  |

### Ethical approval

The study design and protocol have been approved by the ethics committees for human research at our institute (IRB no. R2019-227, R2023-076). Patients’ imaging data were obtained in an opt-out method after their personal information was anonymized in a linkable manner. This study followed an observational design and was conducted following the approved guidelines of the Declaration of Helsinki.

### Imaging Acquisition

A Three-Tesla MRI scanner was used (Discovery MR 750W, GE Healthcare, Milwaukee, WI, USA) with a 24-channel head coil for all MRI examinations. The sequence parameters for the 4D-flow of CCF and intracranial DAVF are as follows: repetition time (TR), 6.02 ms; echo time (TE), 3.03 ms; flip angle, 8 degrees; field of view (FOV), 180 mm; matrix, 256 × 256; and voxel size, 0.703 × 0.703 × 1.0 mm, and that of SAVF as follows: TR, 7.63 ms; TE, 3.05 ms; flip angle, 8 degrees; FOV, 300 mm; matrix, 200 × 200; and voxel size, 1.172 × 1.172 × 2.0 mm. The 3D-velocity encoding (VENC) data were obtained from the 4D-flow MR imaging sequence with 120cm/s for intracranial disease (CCF and DAVF) and 40 cm/s for spinal disease (SAVF). The image range differed from lesions of diseases with a focus on the assumed shunt point. The CS was set for the image range center of CCF and CS-DAVF and the TS was set for the upper one-fourth of the range of TS-DAVF in the axial plane, and the conus medullaris was the center for the image range of SAVF in the sagittal plane. MRI was performed on each subject during normal sinus rhythm and the obtained encoding data was synchronized with the peripheral pulse rate measured at the finger, and a total of 12 phases of the cardiac cycle were reconstructed. Mean acquisition time is 5 minutes per a sequence. Furthermore, we used volumetric data obtained using time of flight (TOF) sequences (TR, 25 ms; TE, 3.1 ms; flip angle, 20 degrees; matrix, 512 × 224; voxel size, 0.391 × 0.391 × 0.5 mm; and 208 slices) for CCF and intracranial DAVF to construct patient-specific geometries of the arteries, and gadolinium-enhanced T1-weighted images were used (TR, 5.28 ms; TE, 2.1 ms; flip angle, 12 degrees; matrix 256 × 192; voxel size, 0.781 × 0.781 × 2.0 mm; and 64 slices) for SAVF.

### Data Analysis

Using the 4D-flow application on a stand-alone 3D volume analyzer workstation (SYNAPSE 3D; FUJIFILM Corporation, Tokyo, Japan), 3D flow images with a cardiac cycle were created from time-resolved VENC data of phase-contrast sequences and anatomical information from TOF sequences for CCF/intracranial DAVF or gadolinium-enhanced T1-weighted images for SAVF. Brain parenchyma and vessels were automatically extracted from the CSF spaces and bone structure using a threshold algorithm at 350–450 of the lower limit thresholds and no upper limit threshold on a TOF sequence to analyze the area of 4D-flow analysis of CCF/intracranial DAVF (Fig. 1 A, B). The target area for a 4D-flow analysis of SAVF was extracted with the same threshold algorithm in 2000–2100 of the lower limit thresholds (Fig. 4 B, C). Mean post-processing time is 10 minutes per a study.

**Fig. 1.**
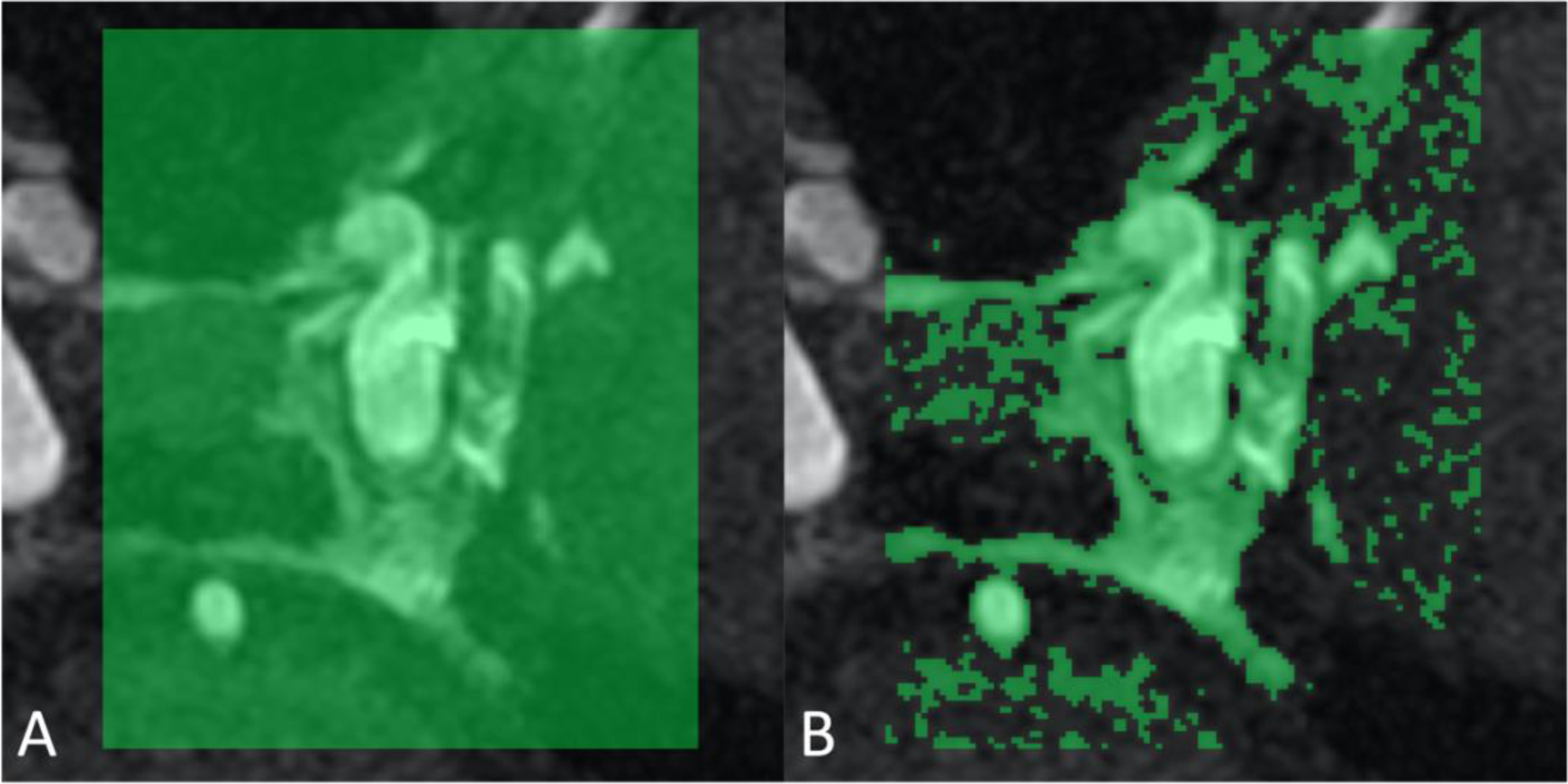
Data analysis using 4D-flow application. The light green area indicates the region of interest for 4D-flow analysis of CCF (A), and the area is automatically extracted from the CSF spaces and bone structure by a threshold algorithm (B).

#### 2D-velocity colormap

Visually detecting the point in which flow velocity increases in the area that included fistulas, we used a 2D velocity colormap that visualizes the velocity with the warm colors that represent high velocity and cool colors that represent low velocity. Color tone intensity of the 2D velocity colormap was adjusted by changing the upper limit threshold of velocity to 80–100 cm/s to more clearly distinguish the difference in colored velocity signal. An accelerated blood flow stream without information of flow direction/vector was obtained by observing three directions, such as axial, sagittal, and coronal, of 2D-velocity colormap and inclining observation axis along the flow direction long axis. The sites where the flow velocity increased were determined as arteries, veins, or venous sinuses based on the cerebral anatomy using the reference TOF images or contrasted T1 images.

#### 2D-flow vector

Furthermore, lesions shown as warm colors and high velocity on 2D-colormap were visualized as 2D-flow vector information. The 2D-flow vector was assessed in three directions and the long axis of flow direction as with the 2D-velocity colormap. The colored arrows on the 2D-flow vector indicate vectors for the cardiac-gated blood flow. The vector direction indicates the blood flow direction, and the color and size indicate the velocity magnitude. The sequence parameters for 2D-velocity colormap are as follows: sampling spaces of 0.5mm and minimum velocity of rendering vector of 10 cm/s.

## Results

Preoperative 4D-flow MRI was performed in 21 of 23 cases, of which the shunt point could be detected in 18 cases, excluding 1 case with TS DAVF and 2 cases with ACF DAVF (Table.1). The results were consistent with those of DSA.

### Representative cases of CCF, DAVF and SAVF. Carotid cavernous fistulas

In 2D-velocity colormap, continuous color image parts were found between the internal carotid artery (ICA) and CS (Fig. 2 B, C). Horizontal blood flow was not visualized in the TOF image (Fig. 2 A) but detected in the 2D-velocity colormap (Fig. 2 B, C, and Supplemental Video S1) and the 2D-flow vector of 4D-flow MRI (Fig. 2 D and Supplemental Video S1) and coronal section of CT-like image of 3D rotational DSA. The 2D-flow vector image showed the blood flow direction from ICA to CS, though that point was confirmed as the shunt point/fistula of CCF. As confirmed on the 2D-velocity colormap and 2D-flow vector, the inflow shunts blood of the CS drained into the drainers and inferior petrosal sinus (Fig.3 D, E and F). One case was treated with coil embolization only, and the other was treated with coil embolization using an intracranial stent placed in the internal carotid artery covered the shunt point. Both cases were archived with complete obliteration of the fistulas with embolizing CS part, and the residual shunt was not detected by DSA and 4D-flow MRI.

**Fig. 2.**
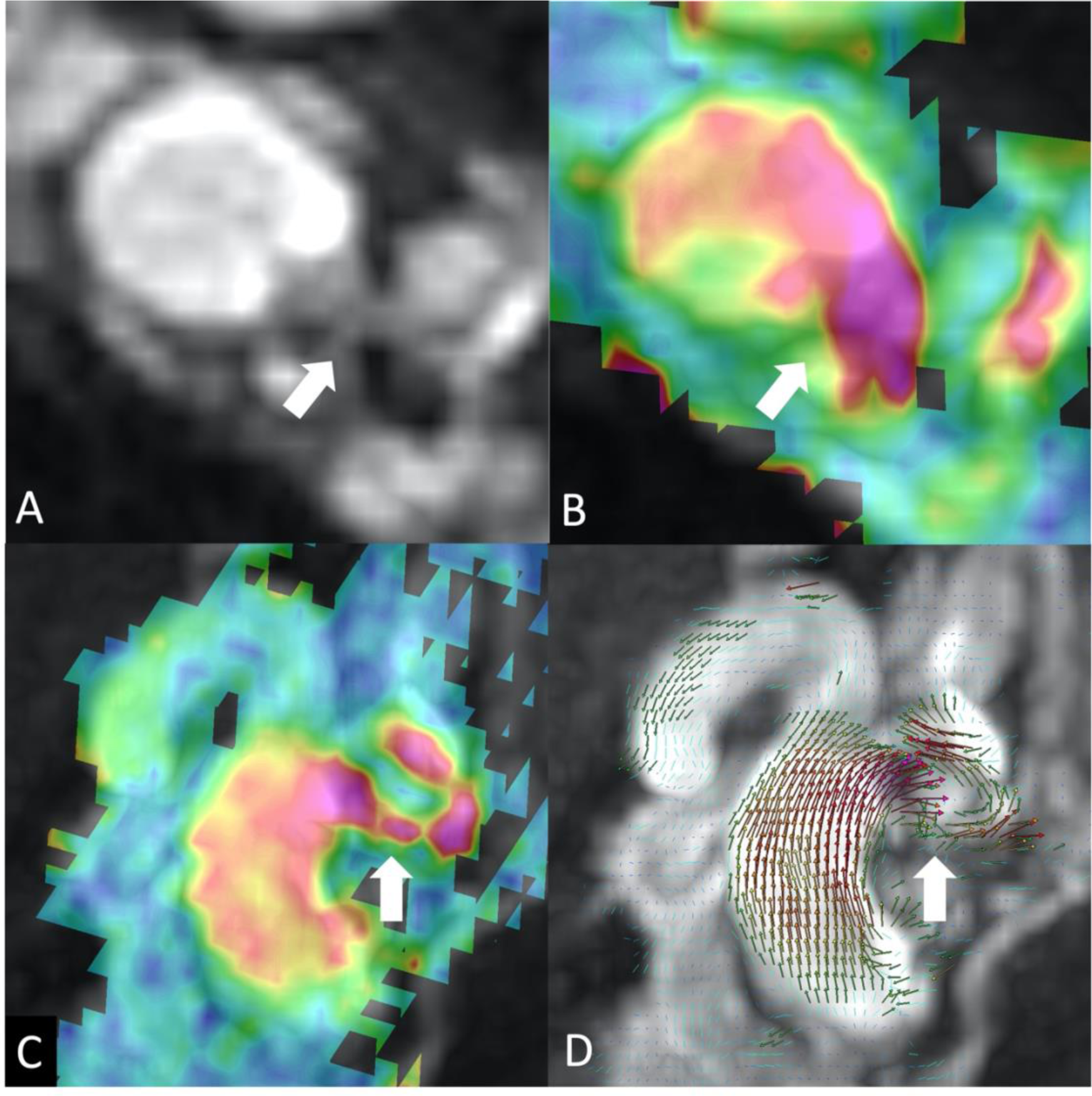
The coronal planes (A, B) and the axial planes (C, D) of the reference TOF images and 2D velocity colormap of CCF (Barrow type A). Horizontal flow of fistula was not visualized on the TOF images (arrow A) but was detected on the 2D-velocity colormap (arrow B). Fistula of CCF displayed in the 2D velocity colormap (arrow C), and 2D-flow vector image showed the direction of blood flow from ICA to CS (arrow D).

**Fig. 3.**
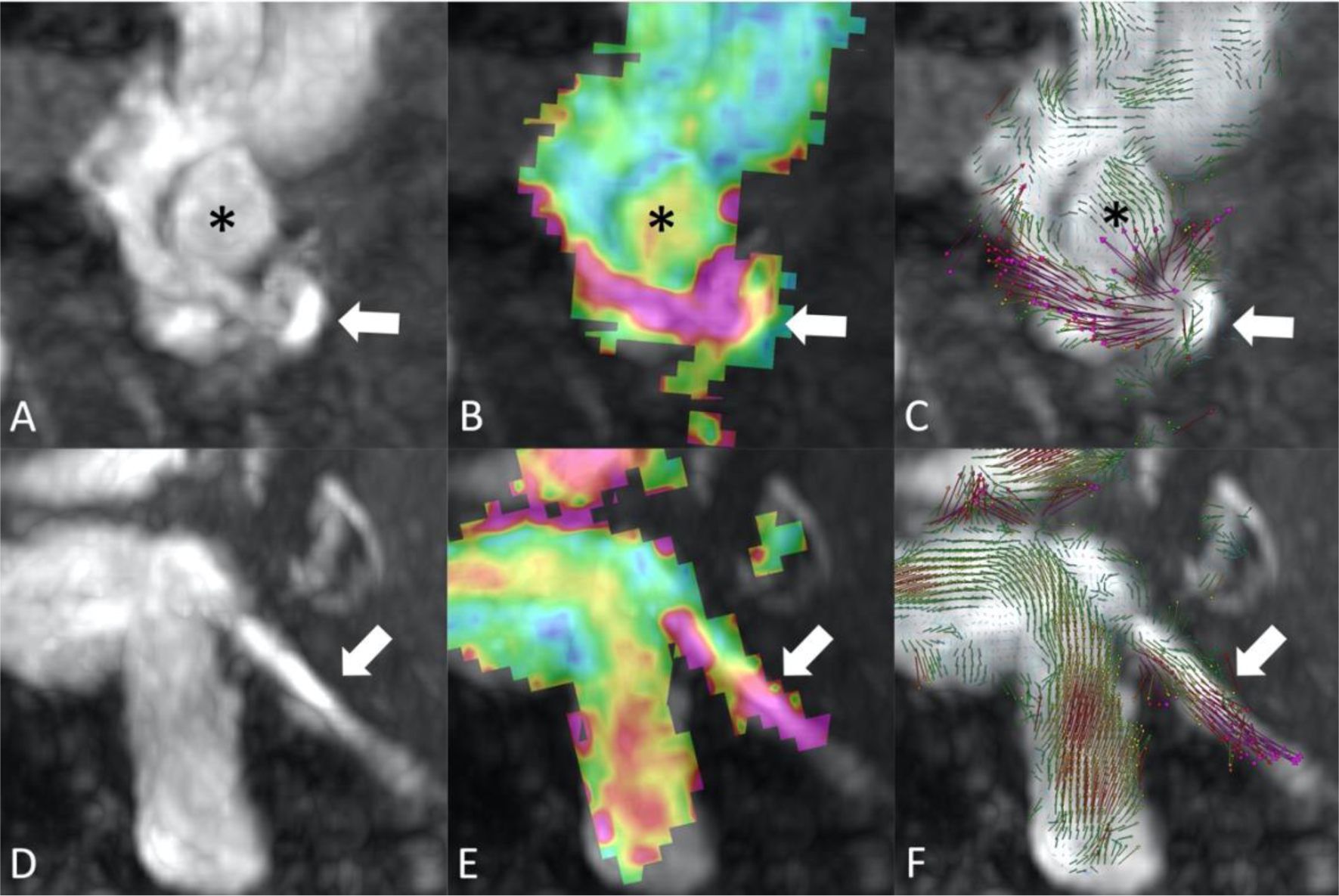
A case of direct CCF (Barrow type B) caused from ruptured aneurysm of persistent primitive trigeminal artery. A to C are same phases of axial planes (asterisk indicate ICA). Blood flow from shunt point (arrow) to cavernous sinus are confirmed in 2D velocity colormap(B) and 2D-flow vector(C). D to F are same phases of sagittal planes. Inferior petrosal sinus as main drainer from cavernous sinus is detected in each images, TOF (D), 2D velocity colormap(E) and 2D-flow vector(F).

### Intracranial dural arteriovenous fistulas

Regarding the five cases of the CS-DAVF, shunt point of a representative case was detected as the point that increase velocity in the 2D velocity colormap in the posteromedial side of the CS (Fig.4 A, B). The other four cases were also detected shunt points around the CS in same manner. In 2D-flow vector images of the representative case above, the ascending pharyngeal artery flowed into the shunt point as a feeder and flowed out to the intercavernous sinus and inferior petrosal sinus as the draining route in the former, and the middle meningeal artery as the main feeder, and the superior petrosal sinus as the main drainer were observed. This case was treated by selective transvenous coil embolization via the inferior petrosal sinus, and completely obliterated (Fig.4 C, D). Two cases of CS-DAVF were received a postoperative 4D-flow MRI assessment, and the result was coincident with postoperative DSA.

**Fig. 4.**
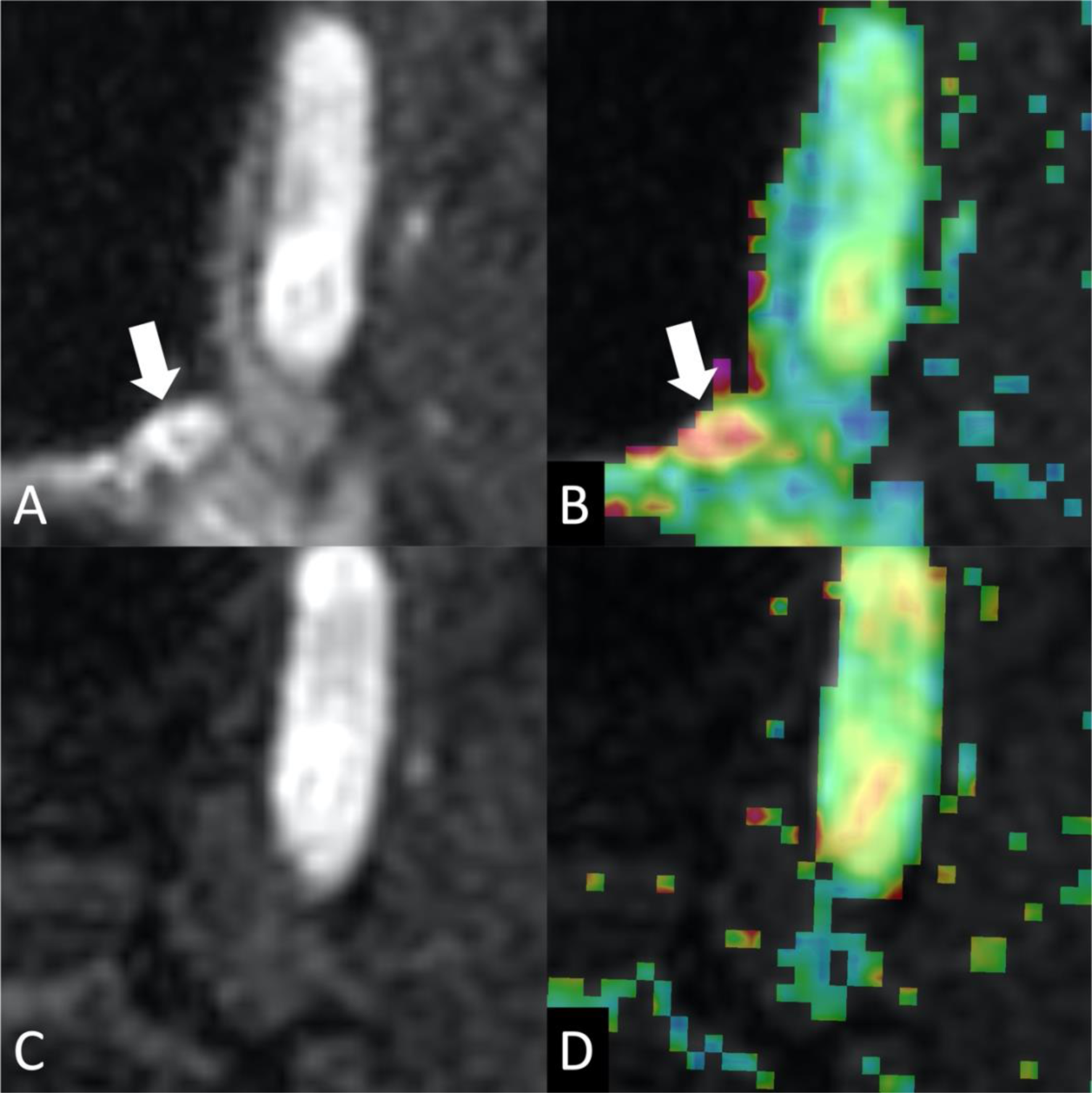
A case of CS-DAVF (Cognard typeⅡa/ Barrow type D). Axial planes of preoperative MRA showed shunt point(arrow) at posteromedial site of CS in TOF(A) and 2D velocity colormap(B). Postoperative TOF image(C) and 4D-flow MRI(D) indicate complete obliteration of fistula.

This study includes ten cases of TS-DAVF. The first representative case of TS-DAVF classified as Cognard type Ⅱa, only underwent MRI, including 4D-flow MRI and DSA, and shunt points were confirmed at the transverse sigmoid junction and jugular foramen (Fig.5). In the second representative case of TS-DAVF, a shunt point was found in the lateral one-third of the TS by 2D-velocity colormap and 2D-flow vector images (Fig.6 A, B and C). The shunting flow was divided into two directions, lateral and medial, and drained into cortical veins of the posterior cerebrum and cerebellum, and the superficial middle cerebral vein via the tentorial sinus (Fig.6 A, B and C). On the first treatment of the second case, the medial component of the fistula was occluded by *n*-BCA, and the residual shunting that flows to the lateral direction was confirmed after the treatment (Fig.6 D, E and F). The third representative case of TS-DAVF had shunt points at the transverse sigmoid junction (Fig.7 A, B) and one-third downstream of the SS (Fig.7 C, D, and Supplemental Video S2). The shunt flow was divided into two streams, ascending in SS and descending in the internal jugular vein from the shunt point, jugular foramen, and prevented normal venous return (Fig.7 D). Shunt flow reduction was achieved by transarterial *n*-BCA embolization in this case. The fourth representative case was right TS-DAVF classified Cognard type Ⅲ, with two times of past endovascular surgical history in other hospitals. The first and second treatments were performed by platinum coil, *n*-BCA, and Onyx through several feeders. Cortical venous reflux worsening was confirmed by DSA after twice endovascular treatments, thereby the third treatment was planned. At third treatment, complete obliteration was achieved by *n*-BCA injection into the branch of the middle meningeal artery, and postoperative DSA showed no residual shunts. Preoperative 4D-flow MRI was performed in this case just before the third treatment, and 2D-velocity colormap could not detect fistula point (Fig.8).

**Fig. 5.**
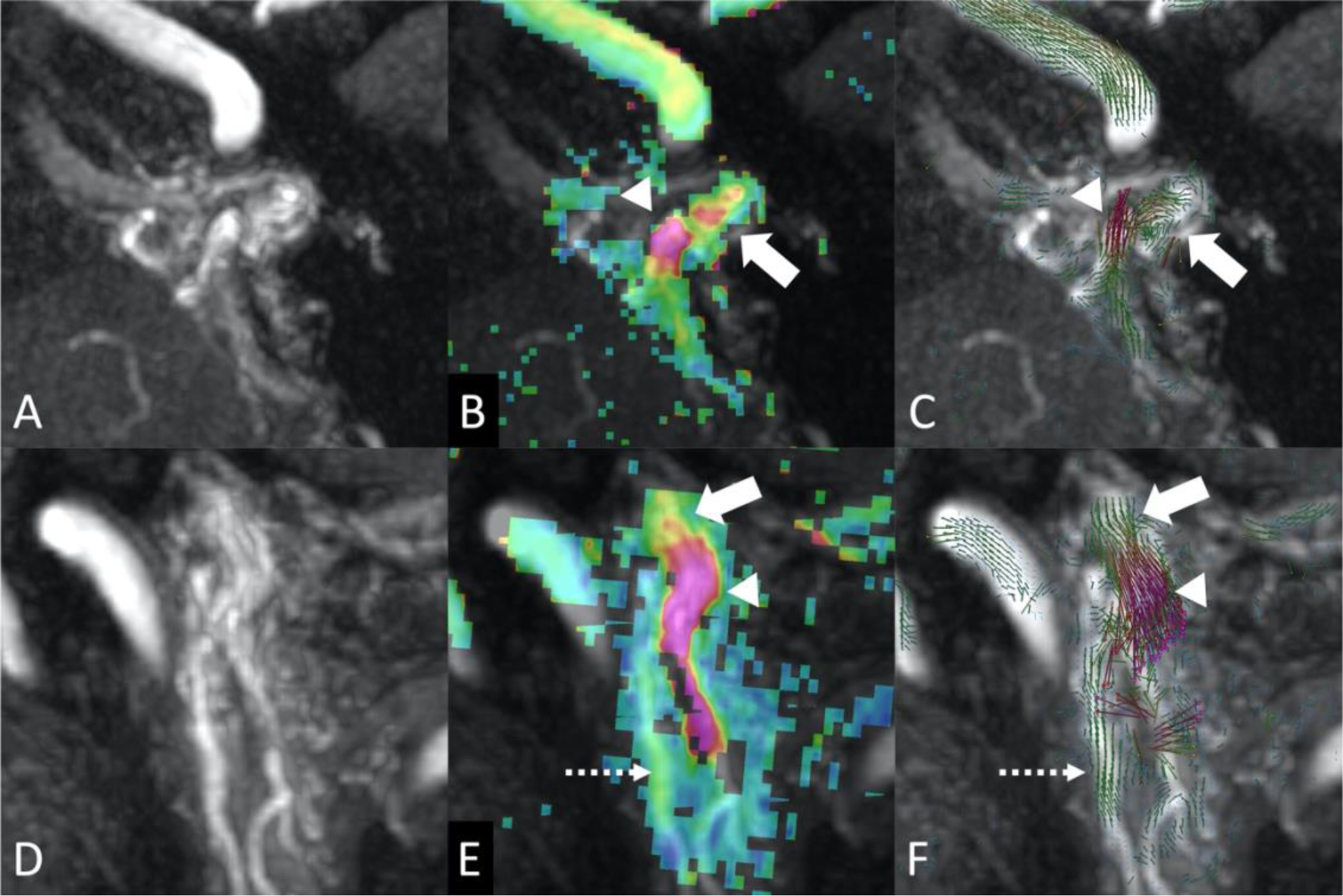
A case of DAVF (Cognard type Ⅱa) which has two fistulas, one is at SS and the other is at transverse sigmoid junction. A to C are axial planes of jugular foramen of TOF(A), 2D velocity colormap(B) and 2D-flow vector(C). Shunt point at jugular foramen (arrow B, C) and a drainer, posterior condylar vein (arrowhead B, C) are detected. D to F are sagittal planes of SS. TOF(D), 2D velocity colormap(E) and 2D-flow vector(F). Shunt point at jugular foramen (arrow E, F), shunt flow in narrowed internal jugular vein and ascending pharyngeal artery as a feeder (dashed arrow E, F) are confirmed.

**Fig. 6.**
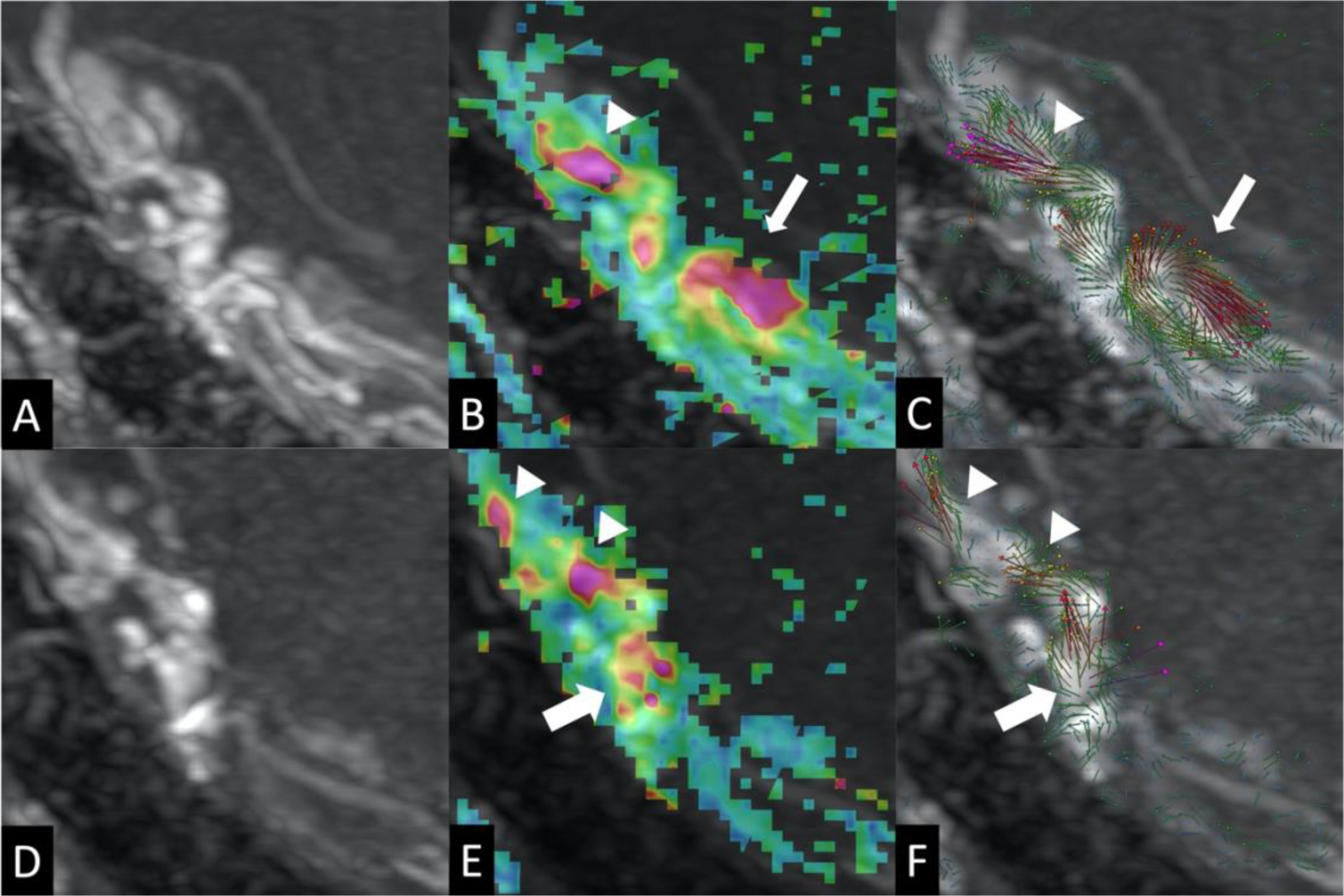
A case of TS-DAVF (Cognard type Ⅱa+b). Axial planes of same phases of TOF(A), 2D velocity colormap(B) and 2D-flow vector(C). TOF(A) shows abnormal hyperintense lesions along lateral side of right TS. The shunting flow were divided into two directions, laterally (arrow B, C) and medially (arrowhead B, C). The medial component of the fistula was occluded by n-BCA on the first treatment, and hyperintense lesion is still found in TOF image (D). The shunting flow to the lateral direction (arrowhead E, F) from the residual fistula (arrow E, F) was confirmed after the treatment.

**Fig. 7.**
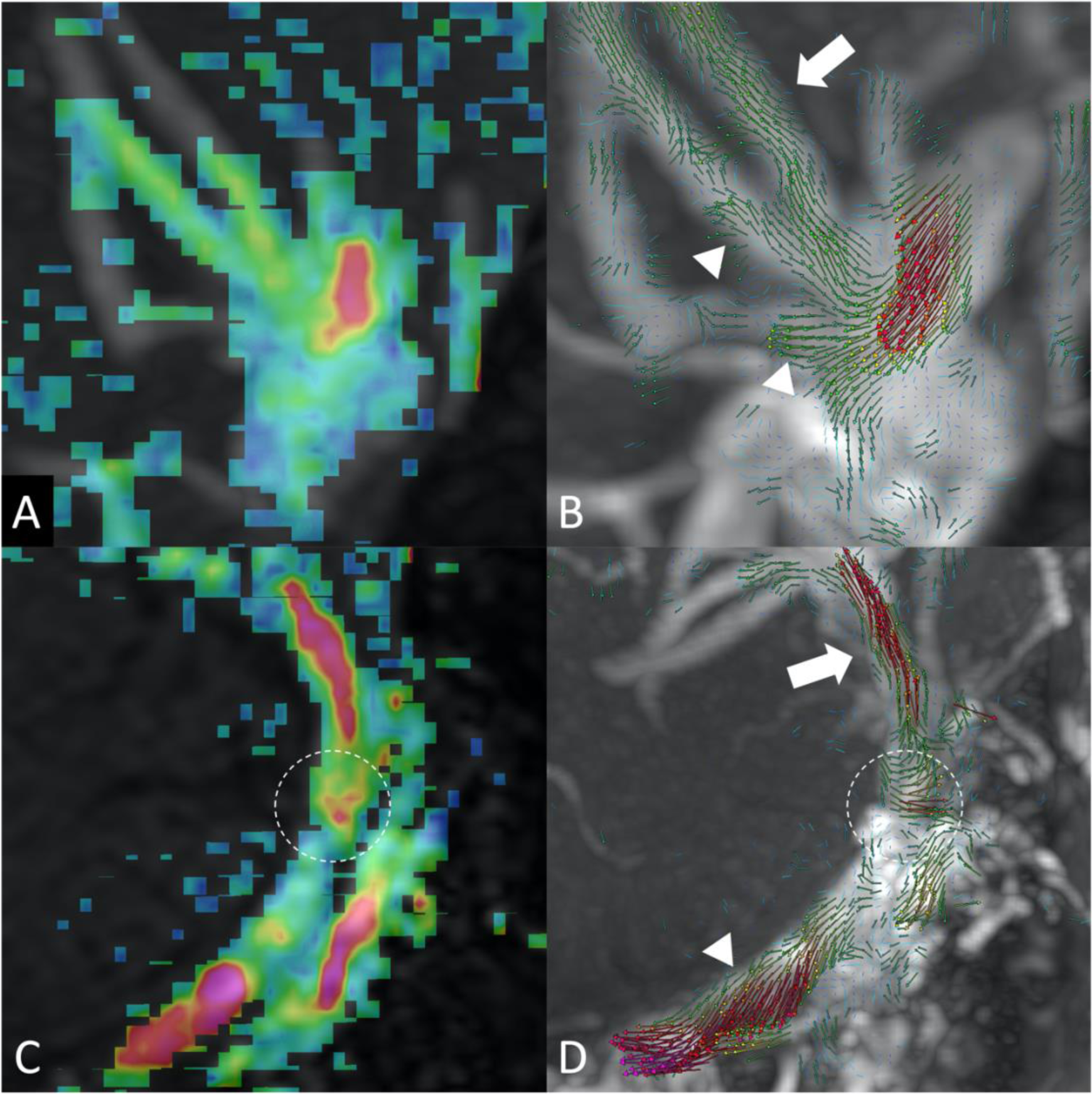
2D velocity colormap of transverse sigmoid junction of DAVF (Cognard typeⅡa+b) visualizes the shunt point as the warm color (A). 2D-flow vector (B) indicates blood flow from shunt point to inferior petrosal sinus (white arrow) and cerebellar veins (arrowheads) as draining veins. 2D velocity colormap of SS DAVF (C) shows a fistula of jugular foramen (dotted circle) and high velocity flow in the SS. The shunt blood flows ascending (white arrow) and descending (arrowhead) within the SS from the shunt point (dotted circle)

**Fig. 8.**
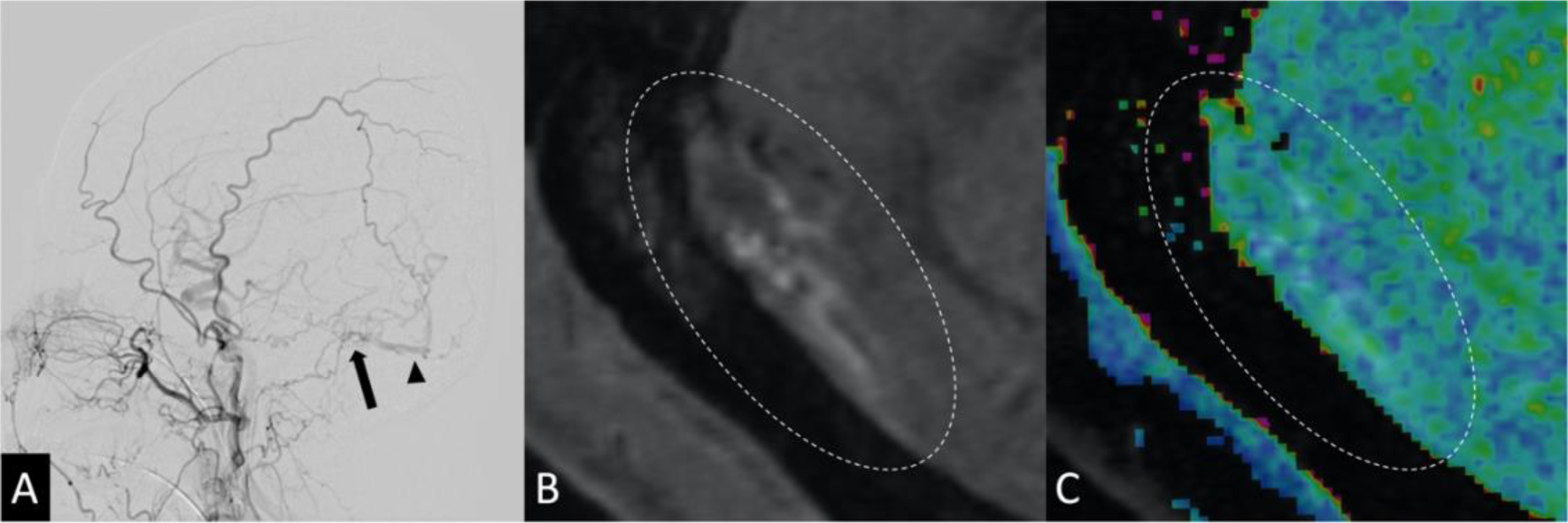
A recurrence case of TS-DAVF (Cognard type Ⅲ). DSA (A), TOF(B) and 2D velocity colormap(C) before third time endovascular surgery. Lateral view of right common carotid angiogram(A) demonstrating shunt point(arrow) and isolated transverse sinus(arrowhead). TOF shows abnormal hyperintense lesions along lateral side of right TS (dotted ellipse B), and 2D velocity colormap has no change in same lesion (dotted ellipse C).

As for two cases of ACF-DAVF, we could not observe the shunt point using 4D-flow MRI because of the artifact of bone structure and air in paranasal sinus. Concerning a case of SR-DAVF and a case of PA-DAVF, shunt detection by 4D-flow MRI were achieved in both cases similarly to other DAVF cases.

In twenty cases of DAVF, preoperative 4D-flow MRI was performed on 18 cases, and shunt points were perceived in 15 cases, excluding TS-DAVF, the case above-described needs three-time treatments, and two cases of ACF-DAVF. Regarding the 15 DAVF cases which shunt point were detected by 4D-flow MRI preoperatively, detected shunt points were all corresponding to the fistulous points identified by DSA at preoperative examination or endovascular operation.

### Spinal arteriovenous fistulas

The case of conus perimedullary AVF presented with neurogenic bladder dysfunction due to a large varix compression, approximately 20 mm at the L1 lumbar vertebral level. The AVF had a complex vascular structure that several feeders, including arteries derived from the left L2 segmental artery, left Th9 segmental artery (the artery of Adamkiewicz), and right L4 segmental artery, were connected through the conus basket (Fig. 9 A). the centered large varix and small varix just beside large varix in 2D-velocity colormap of L1 lumbar vertebral level were visualized as a continuous region that increases flow velocity (Fig. 9 B and Supplemental Video S3). 2D-flow vector images showed that shunt blood from the fistula on small varix flowed through large varix, drained into the main drainage route, right S1 radicular vein, and flowed to the caudal side (Fig. 9 C and Supplemental Video S3). Transarterial coil embolization of one of the feeders from the L2 segmental artery via the dilated radicular artery was performed. Selective angiography with a microcatheter before embolization revealed a fistula point of the feeder on the large varix wall close side to the small varix. The shunt flow from the occluded feeder was hiding in the higher flow from the large varix because the blood flow from this shunt point drains into the right S1 radicular vein same as shunt flow from the small varix, and the blood flow from the small varix was upstream of the flow from the large varix. Findings of 4D-flow MRI and DSA in the main shunt flow and residual fistulas were identical.

**Fig. 9.**
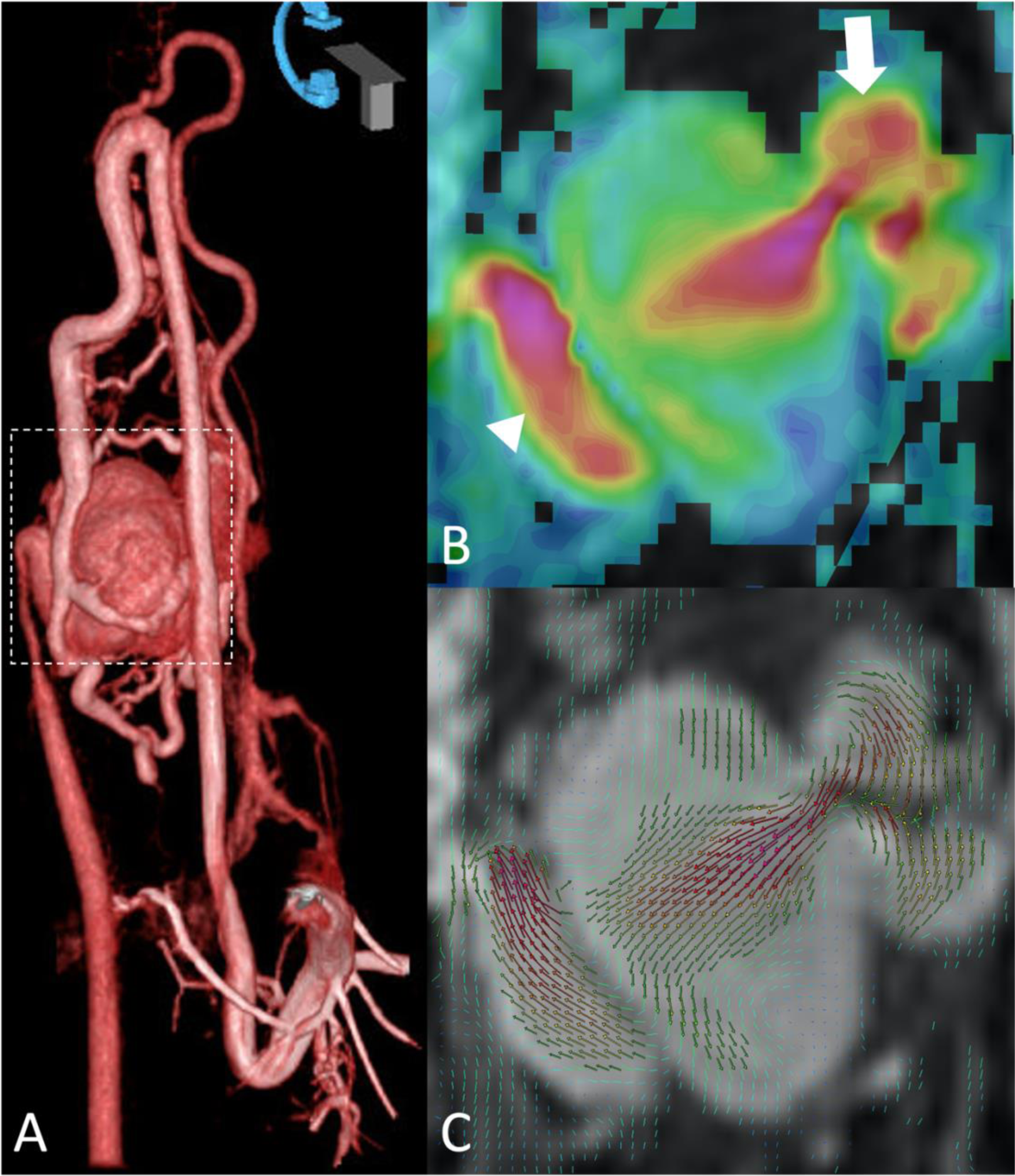
3D rotation angiography by DSA of conus perimedullary AVF (A). The site of coronal plane of 2D velocity colormap (B) and 2D-flow vector (C) are corresponded to dotted square of figure A. 2D velocity map showed blood stream from shunt point(arrow) to the varix centered of the figure, and the main drainage route of a radicular vein(arrowhead). Flow dynamics of vascular complex of spinal AVF is confirmed in 2D-flow vector image(C).

## Discussion

This study showed that 4D-flow MRI allows us to obtain important information about flow dynamics, such as shunt points and flow direction of shunt blood of arteriovenous shunt diseases, including CCF, DAVF, and SAVF. Arteriovenous shunt diseases have the site, called the fistula, where the blood flow velocity is changed due to the pathologic feature of an abnormal connection between arteries and veins ^25–27^.

CCF bring the inflow, the fast blood flow of ICA, into the CS, and the increasing flow velocity in the CS can be captured by 4D-flow MRI. Barrow CCF classification categorizes the CCF with fistulas that have direct communications between the ICA and the CS as type A, and the size of fistulas of Barrow type A was reported as 1–5 mm ^9^. Flow velocity depends on the size of fistulas. No noticeable difference in flow velocity was found between the ICA and the CS through the shunt point in 4D-flow MRI in our CCF cases. The fistula of our CCF cases was also clearly confirmed as a continuous flow of contrast medium by DSA. This is due to the change in flow velocity before and after the shunt point was small in CCF.

Unlike the CCF, the flow velocity of feeders of DAVF and SAVF was slow, especially just the proximal side of the shunt point and the arterial flow adjacent to the shunt point could not be detected by the 2D-flow color map in some cases. Increasing flow velocity was observed in DAVFs except for one retreated case and SAVF cases on the venous side, including the shunt point. The fistula size of the Intracranial DAVF is approximately 30– 200 μm ^28,29^, which is related to the flow velocity changes passing through the shunt point. Bernoulli-Poiseuille equation may explain why blood flow in veins and venous sinuses is increasing, which crosses the shunt point. The narrower the luminal structure which liquid flow through, the faster the flow velocity of the liquid.

Shunt blood flow with different vectors could be confirmed by past evaluation methods, including DSA; however, accurately evaluating the target shunt flow in the overlapping blood vessels is difficult. The most useful point of the 2D-flow vector is the evaluation of such complicated blood flow because of the feature of MRI, which can choose any planes to ideally describe the objective flow. 2D-flow vector facilitates the analysis of flow dynamics in cases with multiple shunt points and cases in which shunt flow goes in multi directions (Fig. 7 and Supplemental Video S2).

TOF MRA was one of the standard examinations for evaluating arteriovenous fistulas. The vascular flow was observed as a hyperintense signal in a dural venous sinus or cortical vein ^30^, and as hyperintense nodules or lesions around the dural venous sinus and cerebral vein. Our study confirmed a part of shunt points where the hyperintensity of TOF and the fistulas by other examination coincided; however, some shunt points were not described on TOF MRI. This is caused by the limitation of the TOF angiographic method. The TOF technique is known to maximally enhance the blood flow when the blood vessel is perpendicular to the plane of imaging ^31^. Therefore, the blood signal is saturated and decreases in a blood vessel that runs parallel in the imaging plane, such as the petrous portion of the ICA or horizontal segment of the middle cerebral artery ^32^. The same condition applies to shunt blood flow, thus shunt flow parallel to the imaging plane is poorly visualized. 4D-flow MRI technique is not imposed directional limitation, on this point detecting shunt blood flow using 4D-flow velocity colormap is superior to the TOF MRI method.

The higher blood flow from the shunt point received by the main feeder of many feeders masks the flow from other shunt points when there are multiple fistulas as in the SAVF of this study. Using 4D-flow MRI to discriminate overlapped shunt flow is difficult, hence assuming shunt points from different perspectives by several evaluation exams is important. Precise fistula detection or sufficient understanding of flow dynamics of arteriovenous shunt disease cannot be achieved by a single measurement. Moreover, the most essential key to analyze the arteriovenous shunt disease is deep comprehension of anatomical vascular structure as fundamental knowledge.

Our results showed that 4D-flow MRI may not detect shunt point of ACF-DAVF. The feeder of the ACF-DAVF is the ethmoid artery^33^, which runs within the ethmoid sinus, contains air, and around the ethmoid bone including the crista galli. We could not observe the shunt point using 4D-flow MRI because of the artifact of air of the ethmoid sinus and bony structure. Because both air and bone show weak signals similar to blood flow, it is difficult to distinguish between them when they are assembled together in a small space.

4D-flow MRI can be performed without contrast media; thus, the exam is available as a non-invasive evaluation at least for intracranial DAVF and CCF. We used contrast-enhanced MRI for the anatomical reference image of SAVF for better visibility; however, we confirmed that non-contrast-enhanced fast imaging employing steady-state acquisition (FIESTA) can be an alternative reference for it. A study reported about the splanchnic flow that they used True FISP, which is a coherent gradient echo sequence same as FIESTA, for imaging arterial and venous vessels for reference of 4D-flow MRI ^34^. Meanwhile, other imaging methods, including contrast-enhanced CT, can be used for reference images to 4D-flow MRI in addition to contrast-enhanced MRI and FIESTA. Pursuing the optimum image conditions for 4D-flow MRI is based on patient invasiveness and practitioner visibility.

Our study has some important limitations. First, the unknown influence of susceptibility artifact to 4D-flow MRI from the platinum coil used for the treatment ^35^. We evaluated five cases that used platinum coils in catheter treatment, the results of 4D-flow MRI and DSA were consistent in both cases with residual shunt and cases in which the shunt had disappeared. In these five cases with platinum coils, 4D-flow MRI was sufficient to detect arteriovenous shunt although more cases are needed to verify the effect of coil. Second, treatment-related noise caused by Onyx was observed in two cases. Liquid materials, such as *n*-BCA and Onyx, were produced with any apparent artifacts in MRI; however, Onyx was visualized as a hypointense lesions ^36^ comparable to the CSF space and bone structure in MRA used for a reference image of 4D-flow MRI. However, casts of the Onyx seem to be a weak blood flow signal as noise on 4D-flow MRI, and the noise can be reduced with the threshold algorithm, which is the same measurement mentioned in the data analysis. Third, arteriovenous shunting diseases, such as CCF and DAVF, are rare diseases; therefore, we could target only a few cases and limited DAVF types for 4D-flow MRI analysis. Our study could not include some types of DAVF, such as DAVF of craniocervical junction, and tentorium. Additionally, there is no validated 4D-flow MRI method for arteriovenous shunting diseases. Optimizing the imaging conditions, especially the image used as the reference and the VENC settings according to types of diseases, 4D-flow MRI is expected to make a more accurate shunting disease diagnosis.

## Conclusions

Detecting fistulas point and analyzing flow dynamics of arteriovenous shunt diseases by 2D-velocity colormap and 2D-flow vector of 4D-flow MRI were found reliable for diagnosis. The diagnosis results by 4D-flow MRI coincide with the diagnosis by DSA except for one case, and we have concluded that the case could not detect the fistula point caused by inappropriate VENC settings. 4D-flow MRI used for arteriovenous shunt disease assessment is a method different from past measurements such as contrast-enhanced CT, contrast-enhanced MRI, TR 3D MRA, and DSA, hence it can complement those examinations.

The present study suggested that 4D-flow MRI is a feasible and useful method based on flow dynamics for preoperative and postoperative arteriovenous shunt disease assessments. 4D-flow MRI is expected to play a different role from past assessment method for evaluating arteriovenous shunt diseases although it requires accumulation of cases to be established as a confirmed method.

## Data Availability

The data that support the findings of this study are openly available

## Acknowledgments

We would like to thank the radiology staff of the Shiga University of Medical Science, particularly Masahiro Yoshimura, and Shinnosuke Hiratsuka. We thank Enago (www.enago.jp) for the English language review. We are grateful to FUJIFILM Corporation for allowing use to use the pre-released version of SYNAPSE 3D workstation.

## Supplemental Material

Videos S1-S3

## Nonstandard Abbreviations and Acronyms

CCF: carotid cavernous fistula
DAVF: dural arteriovenous fistula
SAVF: spinal arteriovenous fistula
4D-flow MRI: four-dimensional-flow magnetic resonance imaging
DSA: digital subtraction angiography
CS: cavernous sinus
TS: transverse sinus
SS: sigmoid sinus
ACF: anterior cranial fossa
SR: sphenoid ridge
PA: petrous apex
VENC: velocity encoding
TOF: time of flight
DSA: digital subtraction angiography
ICA: internal carotid artery

